# Coughs, Colds and “Freshers’ Flu” Survey in the University of Cambridge, 2007-2008

**DOI:** 10.1101/2021.03.31.21251220

**Authors:** Ken TD Eames, Maria L Tang, Edward M Hill, Michael J Tildesley, Jonathan M Read, Matt J Keeling, Julia R Gog

**Affiliations:** Department of Applied Mathematics and Theoretical Physics, Centre for Mathematical Sciences, University of Cambridge, Cambridge, CB3 0WA, UK; Joint UNIversities Pandemic and Epidemiological Research, https://maths.org/juniper/; The Zeeman Institute for Systems Biology & Infectious Disease Epidemiology Research, School of Life Sciences and Mathematics Institute, University of Warwick, Coventry, CV4 7AL, UK; Lancaster Medical School, Lancaster University, Lancaster, UK

## Abstract

Universities provide many opportunities for the spread of infectious respiratory illnesses. Students are brought together into close proximity from all across the world and interact with one another in their accommodation, through lectures and small group teaching and in social settings. The COVID-19 global pandemic has highlighted the need for sufficient data to help determine which of these factors are important for infectious disease transmission in universities and hence control university morbidity as well as community spillover. We describe the data from a previously unpublished self-reported university survey of coughs, colds and flu-like symptoms collected in Cambridge, UK, during winter 2007-2008. The online survey collected information on symptoms and socio-demographic, academic and lifestyle factors. There were 1076 responses, 97% from University of Cambridge students (5.7% of the total university student population), 3% from staff and <1% from other participants, reporting onset of symptoms between September 2007 and March 2008. Undergraduates are seen to report symptoms earlier in the term than postgraduates; differences in reported date of symptoms are also seen between subjects and accommodation types, although these descriptive results could be confounded by survey biases. Despite the historic and exploratory nature of the study, this is one of few recent detailed datasets of flu-like infection in a university context and is especially valuable to share now to improve understanding of potential transmission dynamics in universities during the current COVID-19 pandemic.

## 1 Introduction

There is significant potential for spread of infectious respiratory illnesses at universities. In addition to the annual influenza season, the start of which typically coincides with the start of the academic year for UK universities [1], the current COVID-19 global pandemic has highlighted the need to understand and control high morbidity of infectious disease amongst university students [2]. University students represent a unique relatively closed population with extremely high levels of close social mixing, yet where there is a relatively limited level of enforceable control. Outbreaks in student communities also risk spillover into the general population, including from students travelling home in between terms [3, 4], as well as negatively and significantly impacting the health and academic performance of students [5].

Several recorded outbreaks report high influenza-like illness (ILI) attack rates for students [6, 7, 8]. In the UK, the coined term ‘*freshers’ flu*’ attributes these outbreaks to first-year students who, at the start of the academic year, may be exposed to new infections from the mass migration of students from across the world through socialising. Higher incidence has been observed for meningococcal disease in first-year students compared to higher years [9, 10], and for influenza in undergraduate-aged students compared to older students or faculty staff [7, 6].

There are many possible routes of infection within a university setting. Population density in university accommodation can be high, with many students sharing kitchen or bathroom facilities. As such, students living on campus have been observed to have higher attack rates than those living off campus [7]. Dormitory rooms in particular have been considered to be common settings for likely transmission of respiratory pathogens [11], as well as dormitory common areas and also small classes [12]. Other factors such as catered university accommodation [13], mid-term vacations [8] and specific social calendar events [14] have all seemed to contribute to close-contact infectious disease outbreaks in universities.

Targeted epidemiological data on university students linked with demographic and lifestyle data can help determine the role of different factors impacting the spread of infectious diseases in a university and thus inform suitable preventative and intervention measures. Most available ILI university datasets of this kind to date are from the US [6, 7, 8, 11, 14, 15], few are from recent years [12], and most contain only limited demographic and lifestyle data. Since the start of the COVID-19 pandemic, more datasets have become available from US university COVID-19 studies [16, 17, 18, 19, 20], but universities can differ greatly in structure and student behaviour between countries.

In this paper, we analyse and share the data from a previously unpublished UK university survey of self-reported coughs, colds and flu-like symptoms in Cambridge, gathered between October 2007 and mid-February 2008, during the 2007-2008 influenza season. The online survey had 1076 responses, mainly from students of the University of Cambridge, and surveyed epidemiological as well as demographic, academic, domestic and social information. Although an imperfect exploratory study, it is valuable as one of the few detailed datasets on recent influenza-like infection in a university setting prior to the COVID-19 pandemic, and especially pertinent to share now because of the need to understand and predict transmission within universities in the current COVID-19 pandemic.

## 2 Methods

### 2.1 Data collection

The survey targeted participants who were experiencing or had experienced any kind of self-defined cough, cold or flu-like symptoms. In this paper, we will refer to these as ILI symptoms in a general sense rather than the clinical definition. Participants were recruited by posters advertising the study in the university colleges, departments and other locations around Cambridge frequented by students. Locations were chosen to try to minimise bias in the demographics of the participants recruited, although this could not be guaranteed. The survey was re-advertised and posters were re-applied with new catchphrases when possible over the duration of the survey. The survey was also circulated to email contacts of each college’s undergraduate and postgraduate student bodies and college nurses in order for them to publicise to their own communities. Participants who opted in were entered into a prize draw. No other interactions were had with survey participants.

The first two terms of the 2007/2008 academic year at the University of Cambridge ran from 2 October to 30 November 2007 and 15 January to 14 March 2008, which is when students were expected to be resident in Cambridge. The survey study period started shortly before the first responses were received on 8 October 2007 and ended shortly after the last recorded responses on 12 February 2008 (the exact dates the survey opened and closed were not recorded).

Participants completed an online survey with the date of their symptom onset, demographic information (including age and gender), details of academic study, college, and social and domestic factors. Participants could submit multiple entries recording separate instances of ILI symptoms, and the number of previous times the survey completed was a survey question. The survey had 1076 responses from 1027 participants. The variables are described in Table 1. To preserve non-identifiability, some variables and options were removed or aggregated (see supplementary Text S1). The study was explained and informed consent obtained from all participants before the survey began. The study was approved by the Human Biology Research Ethics Committee of the University of Cambridge (ref. 2007.06). The anonymous and non-identifiable dataset is available to be requested from the corresponding author.

**Table 1:** Description of variables obtained from the self-reported survey, in the order they were presented in the survey. The data is given in the supplementary information.

| Variable name | Description | Type | Missing values |
| --- | --- | --- | --- |
| submission_time | Date and time of survey submission | Datetime | 0 |
| age | Age band: '<20', '20–24', '25–29', '30–34', '35–39', '40–44', '45–49', '>49' | Categorical | 2 |
| gender | 'Male', 'Female' | Categorical (indicator) | 3 |
| college | Choice between all University of Cambridge colleges, or 'none' | Categorical | 3 |
| academic_status | 'Undergraduate', 'Postgraduate', 'Support staff', 'Academic staff', 'none' | Categorical | 3 |
| subject | If academic status is undergraduate: 'Arts and Humanities', 'Medicine', 'Natural Sciences', 'Social Sciences', 'Other Sciences' (defined in supplementary Text S1); else duplicate of academic status variable | Categorical | 2 |
| year_of_course | '1', '2', '3', '4', 'College affiliation', 'No College affiliation', 'College based', 'Department based'. In this paper, postgraduates are aggregated into one category for this variable. | Categorical | 134 |
| symptom_onset_date | Symptom onset date | Datetime | 0 (compulsory) |
| symptom_duration | Duration of symptoms in days: '1', '2', '3', '4', '5', '6', '7', '>7', or 'not yet over' | Categorical | 10 |
| previous | Number of times you previously completed this survey: '0', '1', '2' | Categorical | 2 |
| accommodation | Type of accommodation: 'Halls', 'College house', 'Rented house', 'Own home', 'Other living' | Categorical | 6 |
| kitchen | Number of other people who regularly use your kitchen: '0', '1', '2', '3', '4', '5', '6', '7', '8', '9', '10', '>10' | Categorical | 19 |
| lectures | Hours in lectures/practicals in the 7 days before you got ill: '0', '1', '2-5', '5-10', '10-15', '15-20', '>20' | Categorical | 16 |
| supervisions | Hours in supervisions in the 7 days before you got ill: '0', '1', '2', '3', '4', '5', '6', '>6' | Categorical | 33 |
| evenings | Number of evenings out in the 7 days before you got ill | Numeric | 19 |
| facebook | Number of University of Cambridge Facebook friends | Numeric (originally free text) | 144 |
| identify | Do you think you could identify the person who infected you? | Categorical (indicator) | 17 |
| behaviour_changed | Whether behaviour changed while ill | Categorical (indicator) | 19 |
| behavioural_changes | Which behavioural changes occurred while ill: 'Missed supervisions', 'Missed work', 'Missed meals', 'Missed social activities', 'Stayed in bed', 'Missed lectures', | Multiple choice | 339 |

### 2.2 Statistical analysis

We fitted a Cox proportional hazards model on the days between the start of the first term and date of symptom onset, and chose models based on Akaike’s information criterion (AIC) [21]. We used participants with a symptom onset date during the first term from 2 October – 30 November 2007 when students were resident in Cambridge. We fit models with combinations of all variables in Table 1, excluding predictors about post-symptom-onset factors: duration of symptoms, whether participants thought they could retrospectively identify who infected them, whether behaviour changed whilst ill and the behavioural changes experienced. We also excluded number of Cambridge Facebook friends, due to the amount of missing values. Removing participants with missing values in any of the factors under investigation gave 735 participants. We used midpoints of the categorical intervals, or endpoints for half-open intervals, for interpretability. For type of accommodation, we aggregated the categories “Own home” and “Other”, which had fewer than 10 participants, with “Rented house”. All non-student options were also aggregated for academic status, year of course and subject.

The proportional hazards assumption was checked by the Grambsch-Therneau proportional hazards assumption test [22]. Proportional hazards models were fitted in *R* with the *survival* package [23], and proportional hazards models with explicit time-dependent co-efficients with the *timereg* package [24].

We also investigate how the risk ratio of different subpopulations depends on the unknown reporting rates, the probability that someone who experiences symptoms reports them in our survey. Subpopulation sizes in Cambridge were taken from the academic year 2007-2008 [25]. Further, we approximate the risk ratios using subpopulation survey response rates from other surveys of university students. This assumes that survey response rates do not depend on whether someone experiences symptoms or not, although we acknowledge the limitations of this (see Discussion). The surveys chosen were limited to those with available within-group survey response rates and chosen to have university student populations and the subject of ILIs where possible (Table S1), in an attempt to give comparable response rates to our survey’s unknown rates.

## 3 Results

### 3.1 Study population

The survey had 1076 responses, mainly from University of Cambridge students (97%), but also university staff (3%) and other participants (<1%) (Figure 1a). The student participants make up 5.7% of the university population (6.2% of the undergraduate population and 4.6% of the postgraduate population) in the 2007-2008 academic year [25]. The majority of survey participants were younger than 25 (Figure 1b) and the survey over-sampled female students compared to the student population (Figure 1d). The distribution of students among colleges in the survey also differed from that of the university (Figure 1c).

**Figure 1:**
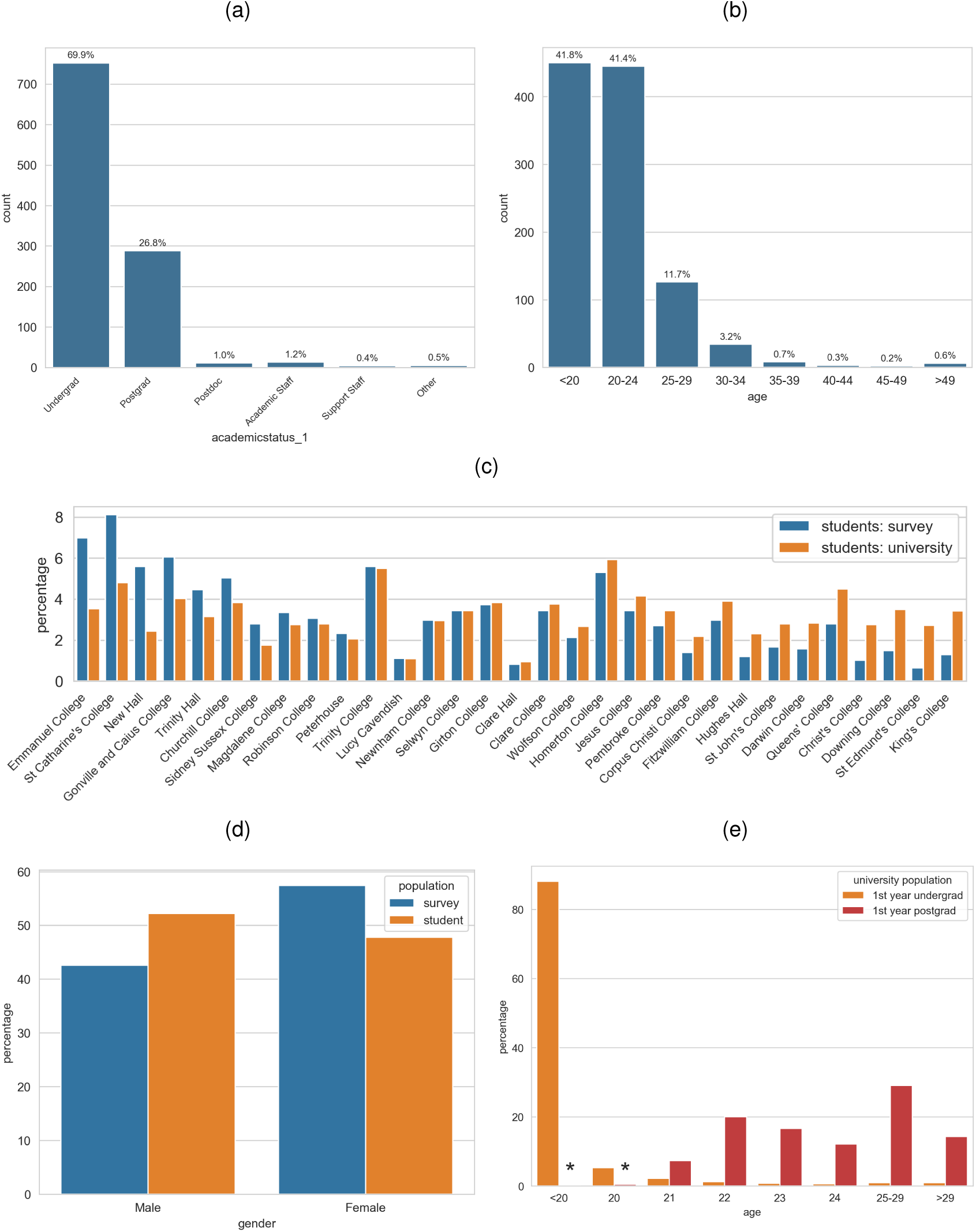
Distribution of participants in the survey by (a) academic status and (b) age. Distribution of students in the survey compared to the student population by (c) college (colleges ordered by most over-represented to most under-represented), and (d) gender. (e) Age distribution of first-year undergraduates and postgraduates admitted to the University of Cambridge in October 2007. *First-year postgraduates aged under 21 total 0.5%. Student population statistics are of full-time undergraduate and postgraduate students in the academic year 2007-2008 from [25].

For undergraduates, the survey over-sampled first-years and under-sampled second and third years, which was also reflected in the undergraduate age distribution (Figure S1) as most first-year undergraduates were aged under 20 (Figure 1e). Undergraduates studying Natural Sciences were also oversampled (Figure S1d).

Postgraduates were under-sampled, compared to undergraduates, but the sample postgraduate age distribution was similar to that of the university postgraduate population (Figure S1). These differences could reflect non-uniform illness incidence between groups, but could also be confounded by sampling biases, such as from the location of advertisements or differences in word-of-mouth advertisement.

### 3.2 Population-level submission summary

In the rest of this results section, the survey output is described as raw data without comparison to the student population, so differences in reported ILI could be a result of sampling biases. The survey ran over the first two terms of the 2007-2008 academic year and gathered 1076 responses between 8 October 2007 and 12 February 2008 (Figure 2a). We observed separate waves during the two terms, with the first wave significantly bigger, and participants also retrospectively reported illness from September and the Christmas holidays (Figure 2b). 90% of participants answered the survey within four weeks of their reported symptom onset date, with 40% answering within a week (Figure S2).

**Figure 2:**
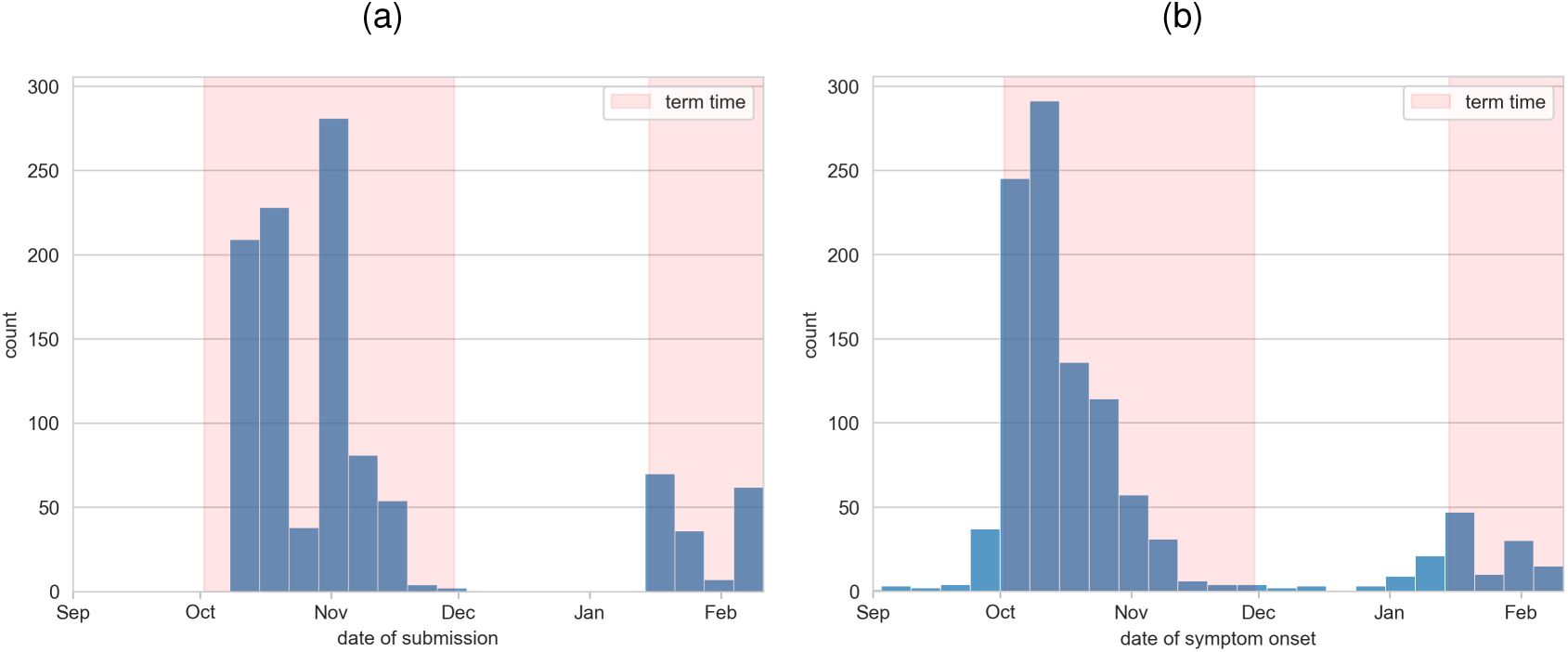
Weekly distribution of (a) survey submission dates, and (b) reported symptom onset date. University of Cambridge academic terms are highlighted in orange.

### 3.3 Demographic factors

There was little difference in timing of reported illness onset between male and female participants (Figure S3a S3b). However, the peak number of undergraduates reporting symptoms in this survey was more than double the postgraduate peak for both waves (Figure 3a). The first wave was also seen to be faster in the undergraduate population compared to the postgraduate population (Figure 3b), but the pattern was less clear for the smaller second term wave. Although the total number of first-year undergraduates with symptoms was larger, there was no evidence that first-year undergraduates became ill before other years, but fourth-year undergraduates were seen to become ill later than undergraduates in other years, at a similar rate to postgraduates (Figures 3c 3d). The corresponding effect is seen with age in Figures 3e and3f, as most students under 20 were first-year undergraduates and most over 24 were postgraduates (as seen in the relationship between age and year of course in Figure 1e).

**Figure 3:**
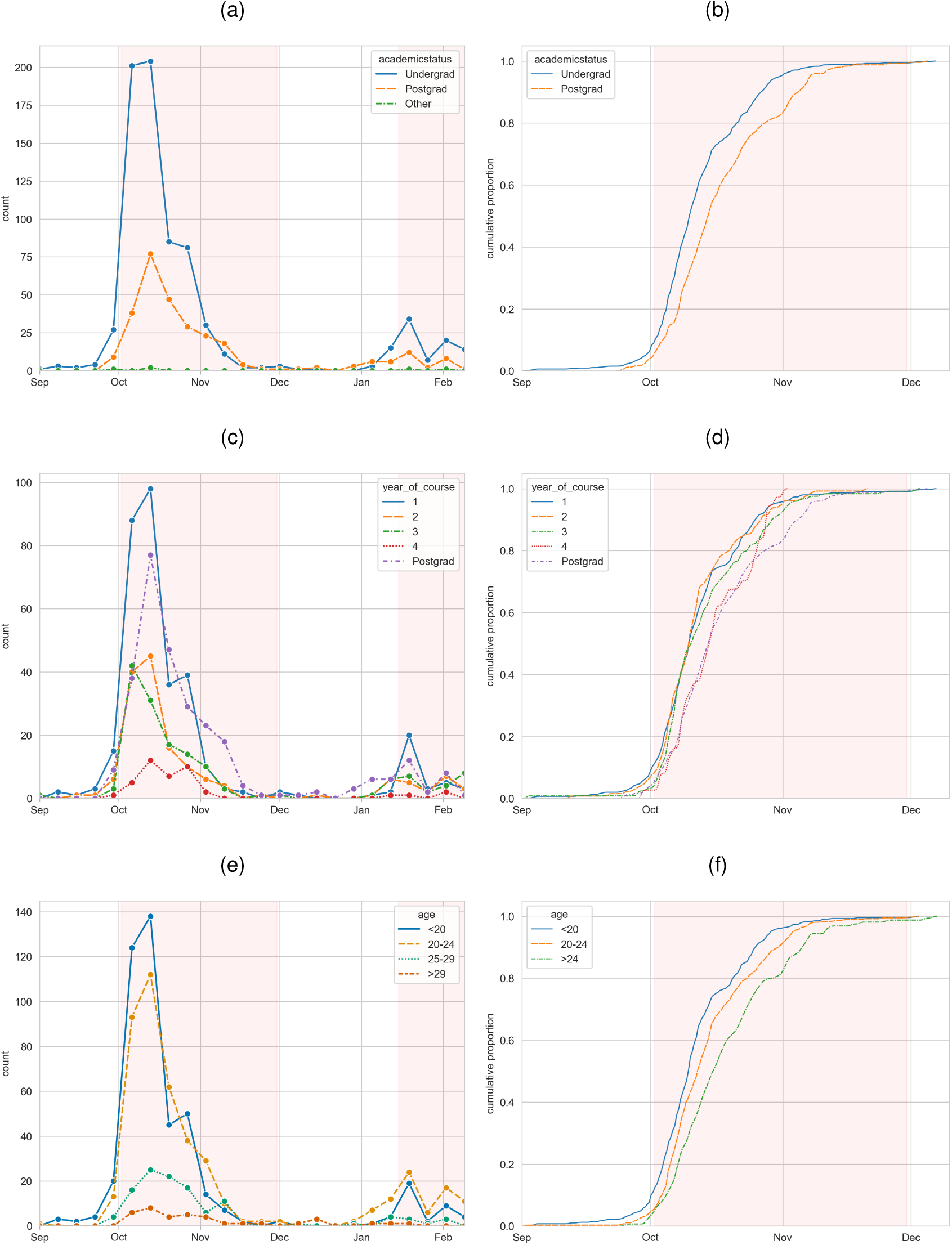
Distribution of weekly reported cases and cumulative distribution of reported cases over the first wave by (a, b) academic status, (c, d) undergraduate year of course and (e, f) age. University of Cambridge academic terms are highlighted in orange.

### 3.4 Academic factors

Out of undergraduate survey participants, the modal response for subject of study was Natural Sciences (Figure S1d), the undergraduate course with the largest cohort [25]. Natural Sciences students were heavily over-sampled, and students studying Medicine and Other Sciences were also slightly over-sampled. This bias could be due to the relevance of the study to these subjects and hence stronger motivation of these students to contribute to the survey. There is some evidence during the first wave of an earlier infection peak in undergraduate students studying Medicine and a later postgraduate peak compared to all other undergraduate subjects (Figures 4a 4b), but this could be confounded by sampling biases (including word-of-mouth advertisement between students on the same course).

**Figure 4:**
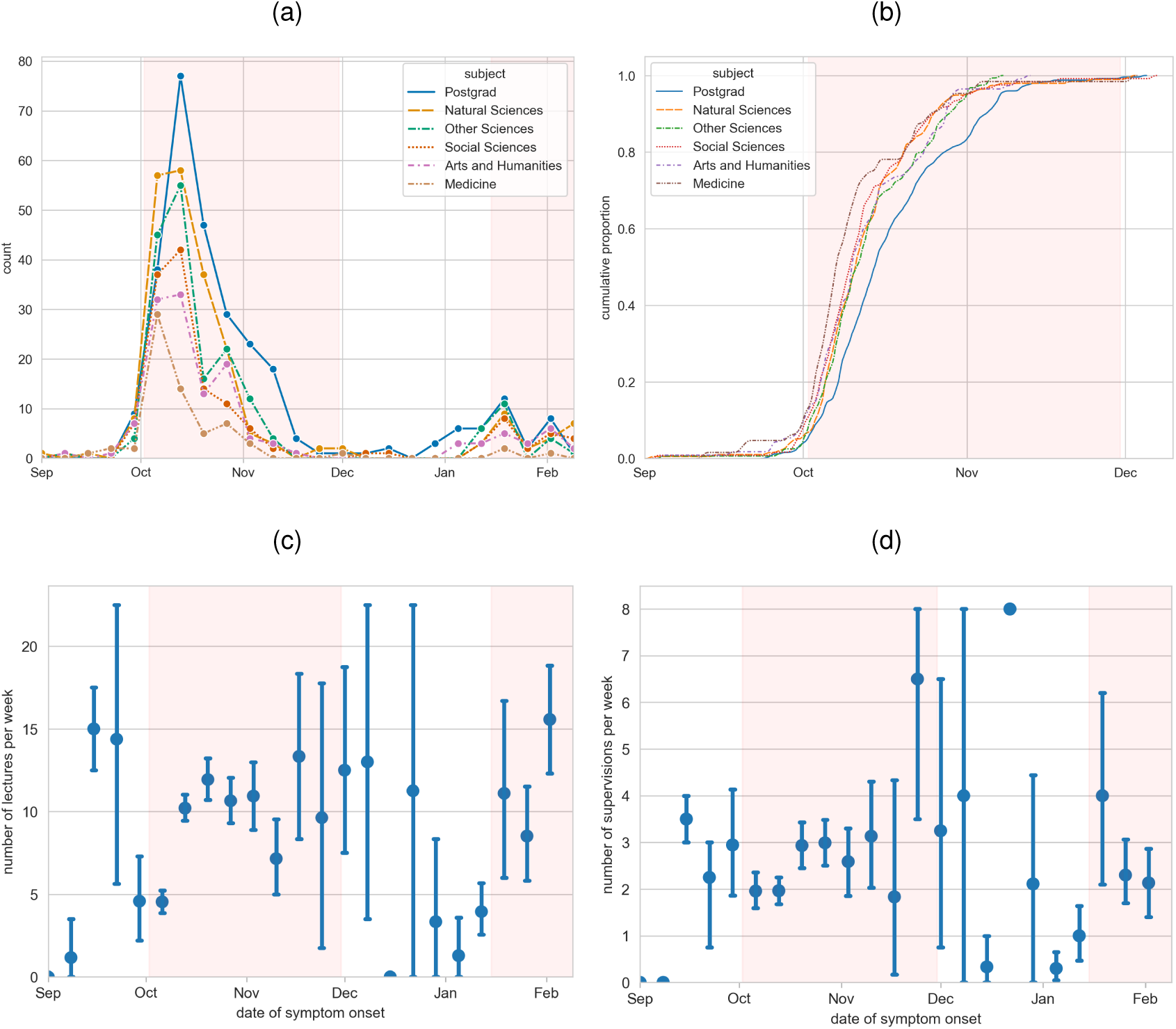
(a) Distribution of weekly reported cases by undergraduate subject or postgraduate status. (b) Cumulative distribution of reported cases over the first wave by undergraduate subject or postgraduate status. (c) Mean number of hours of reported lectures/practicals in the seven days previous to illness, distributed by date of symptom onset. (d) Mean number of hours of reported supervisions in the seven days previous to illness, distributed by date of symptom onset. Error bars show standard error. Midpoints of the categorical intervals are used, or endpoints for half-open intervals. University of Cambridge academic terms are highlighted in orange.

The number of hours of lectures, practicals and supervisions per week can vary greatly between courses and years, as well as individual choices. The size of lecture audiences also varies from a few hundred for courses with compulsory modules and large year groups, to less than ten for courses with many optional modules. Supervisions consist of very small-scale teaching, usually with one to three students per supervisor. The number of lectures and supervisions per week throughout term will vary between courses and years, but lectures tend to start halfway into the first week of term, whereas supervisions in the first term tend to start a few weeks later. Figures 4c and 4d show the relationship between the date of symptom onset and the hours of lectures and supervisions in the seven days beforehand, with no obvious trend and large variances during periods of low total reported cases in September and from mid-November. Some participants with a symptom onset date during the holidays report non-zero hours of lectures and supervisions which may suggest a misunderstanding, such as mistakenly reporting lectures and supervisions from the seven days previous to submitting the survey or from the last seven days of the previous term.

### 3.5 Domestic factors

The University of Cambridge has a collegiate system which is shared by only a few other universities in the UK. In Cambridge, students are generally grouped by college for accommodation and, depending on the course, supervisions (small-scale teaching). Each college offers accommodation, mainly to undergraduates but also postgraduates and some staff, their own centralised catering, and holds social events for college members. Our survey had participants from each of the 31 colleges of the University of Cambridge 1c). There is some evidence of differences in peak height and timing by college (Figure S4), with most first wave peaks occurring before mid-October, but this could be confounded by sampling biases.

Undergraduates are usually expected to live in college accommodation for most of the duration of their degree, which may be on the main college site or elsewhere in the city. Postgraduates are often offered college accommodation for at least the first few years of their degree, which is likely to be off the main college site. These relationships are seen in our survey data (Figure 5a). The majority of reported cases were in halls and the first wave seems to be very slightly earlier in halls than other types of accommodation (Figures 5b and 5c). There is little evidence of a relationship between number of people sharing kitchens with date of symptom onset in term time (Figure S5b). Between terms, most undergraduate students do not reside in their term-time accommodation.

**Figure 5:**
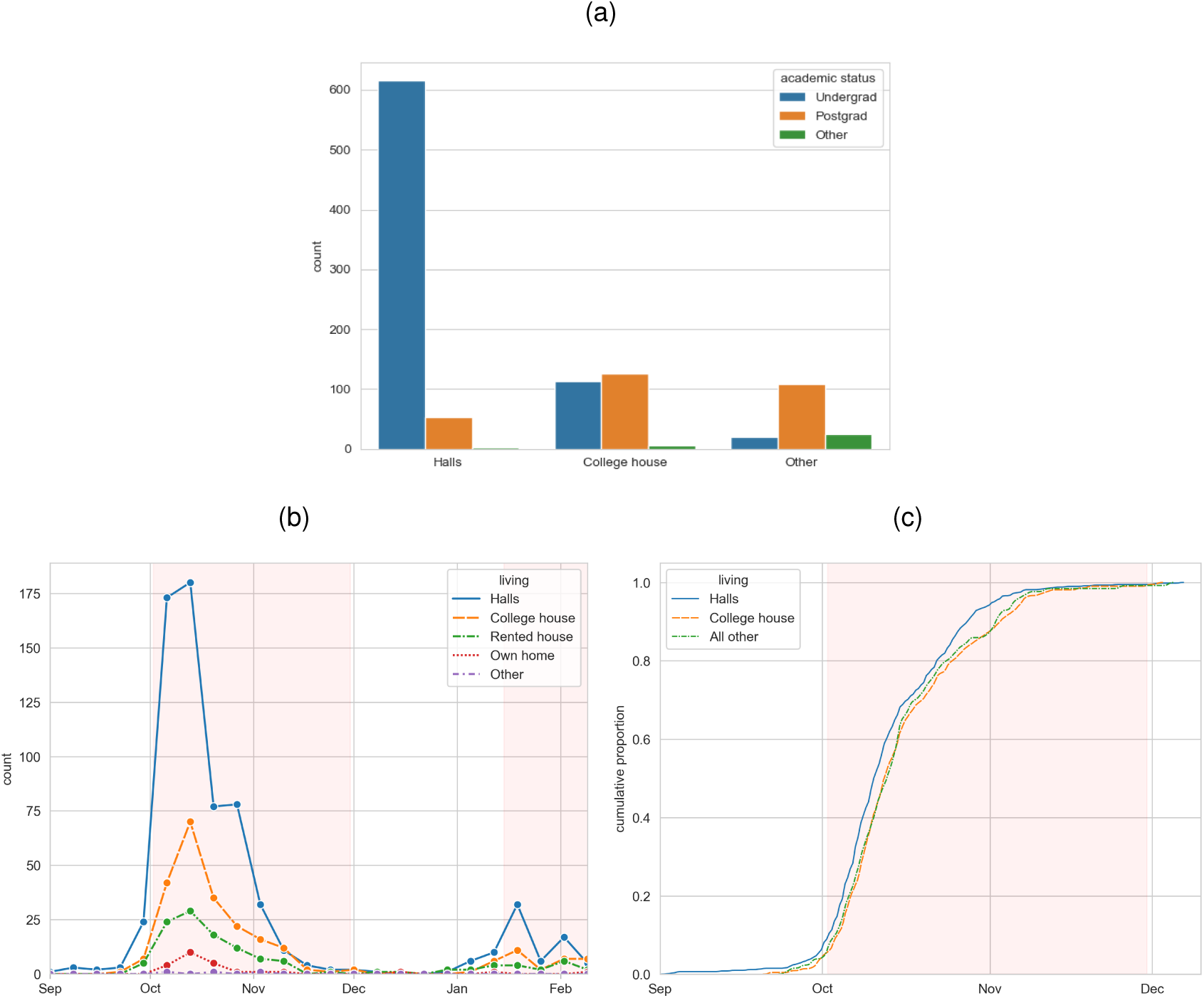
(a) Distribution of reported cases by type of accommodation and academic status. (b) Distribution of weekly reported cases by type of accommodation. (c) Cumulative distribution of reported cases over the first wave by type of accommodation. University of Cambridge academic terms are highlighted in orange.

### 3.6 Social factors

In the first term, there is some indication that students reporting symptoms earlier in the term also reported more evenings out in the week previous to their symptom onset (Figure 6b). However, it might be expected that students go on more evenings out at the beginning of the first term when there are many social events for ‘Freshers’ week’, and evenings out may reduce over the term as workload increases. The increase in evenings out reported in the last seven days of term might also be expected as part of an increase in socialising at the end of term. A definition of ‘evenings out’ was not specified. Outside of term time, university students might display different social behaviour compared with the term time behaviour reported in the survey, so any analysis should consider time in and out of term separately.

**Figure 6:**
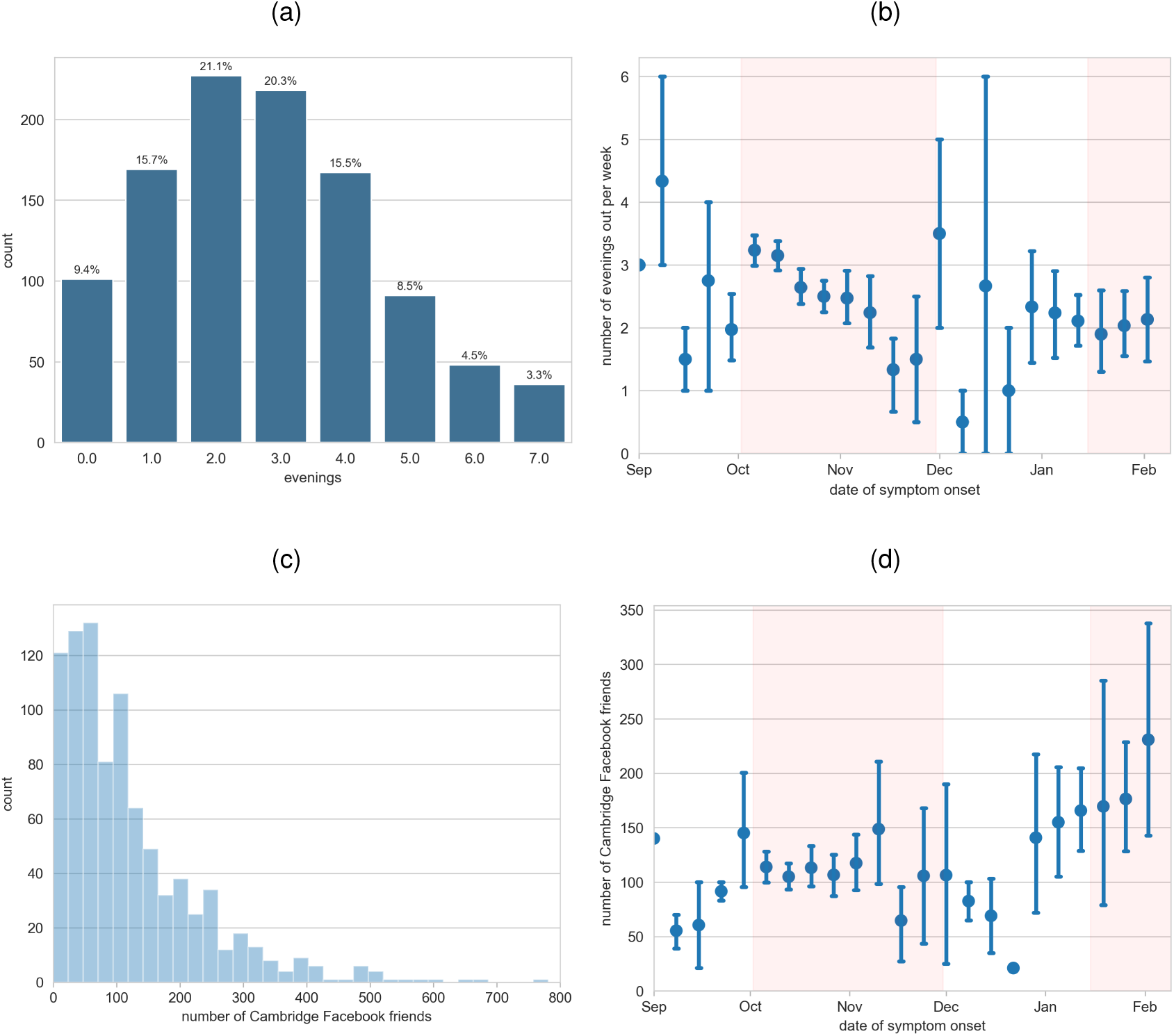
Social factors: (a) Distribution of reported number of evenings out in the seven days previous to illness. (b) Reported number of evenings out in the seven days previous to illness distributed by date of symptom onset. (c) Distribution of reported Cambridge Facebook friends. (d) Reported Cambridge Facebook friends distributed by date of symptom onset. Free text values for Cambridge Facebook friends were only retained if an integer was given. University of Cambridge academic terms are highlighted in orange.

Facebook was a very popular social media platform at the time of survey for university students - a number of surveys in various universities in the US have found that very high proportions of the student population had a Facebook account (94% in 2007 [26], 82% of undergraduates in 2008 [27]). In our survey, 86% of student participants answered the question on Facebook friends at the University of Cambridge with an exact or estimated number, suggesting they had a Facebook account. In 2007, the number of Facebook friends at the same university as you was clearly displayed on your profile. Participants who answered with an exact number reported between 0 and 781 Cambridge Facebook friends, with an overall mean of 119 (s.d. = 110) (Figure 6c). Undergraduates had a mean of 132 (s.d. = 115) and postgraduates had fewer Cambridge Facebook friends on average than undergraduates, with a mean of 93 (s.d. = 93). Other surveys of undergraduates in the US report a similar positive skew in the distribution of all Facebook friends (not limited to the same university) and higher means of 150-200 in 2006 (before Facebook opened to everyone without a school or university affiliation) [28] and 395 in 2008 [29].

Number of Facebook friends has been seen to significantly and positively relate to the rate of upper respiratory infections in university students [30]. It has also been thought to be an indicator of sociability (since seen to be positively related with extroversion [31] and negatively related to anxiety about face-to-face communication [32]) which could correspond to in-person interactions in which infection could spread. In our survey, looking in particular at the number of Facebook friends also attending the University of Cambridge could hence correlate with number of face-to-face interactions within the university community. Figure 6d shows the distribution of Cambridge Facebook friends by date of symptom onset, with no obvious trend across the first term. After Christmas, there is some indication of a positive trend in Cambridge Facebook friends with symptom onset date, but this may reflect a general trend of adding more friends throughout the academic year, particularly to keep in contact with over the vacation.

### 3.7 Symptoms and infection

Most participants reported showing symptoms at the time of answering the survey, but for those whose symptoms had ended, the modal duration of symptoms was more than seven days (Figure 7a). The majority of participants (95%) reported only one incidence of illness in the survey, and less than 5% completed the survey at least twice (Figure 7b).

**Figure 7:**
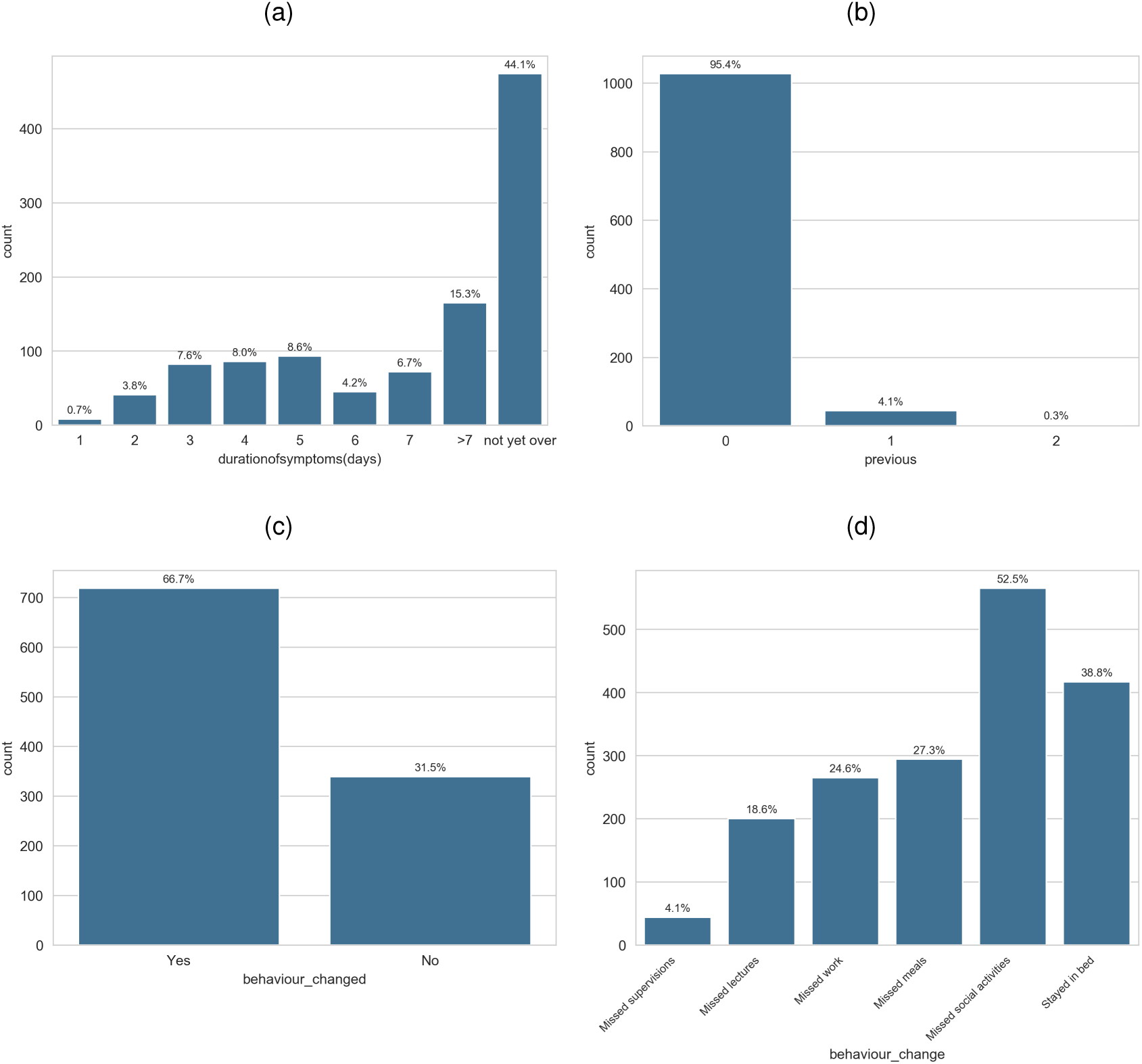
Distribution of survey answers for (a) duration of symptoms in days, (b) previous occurrences of illness, (c) whether behaviour changes were experienced, and (d) if so, which ones (multiple choice).

Participants also reported whether they experienced behavioural changes while ill and if so, which ones (Figures 7c, 7d). 67% reported a change in behaviour as a result of their symptoms. The most common behavioural changes were missing social activities (reported by 53% of all participants) and staying in bed (39%). Participants also missed meals (27%), missed work (24%) and missed lectures (19%), but few missed supervisions (4%).

23% of participants thought they could identify the person who infected them. Whether they were correct would be confounded by many personality and behavioural traits, but a crude method for targeted backwards contact tracing could still be useful to identify likely settings or relationships for transmission.

## 4 Statistical analysis

### 4.1 Proportional hazards model

Given the available data, the three most preferred proportional hazards models by AIC agreed that the number of lectures and supervisions in the week before symptom onset significantly decreased the risk of symptom onset by a factor of 6% per lecture and 4-5% per supervision (Table 2). Third-year undergraduates were also consistently found to have lower risk than first-year undergraduates by around 30%, whereas second and fourth-year undergraduates were not significantly different at the 0.05 level.

**Table 2:**
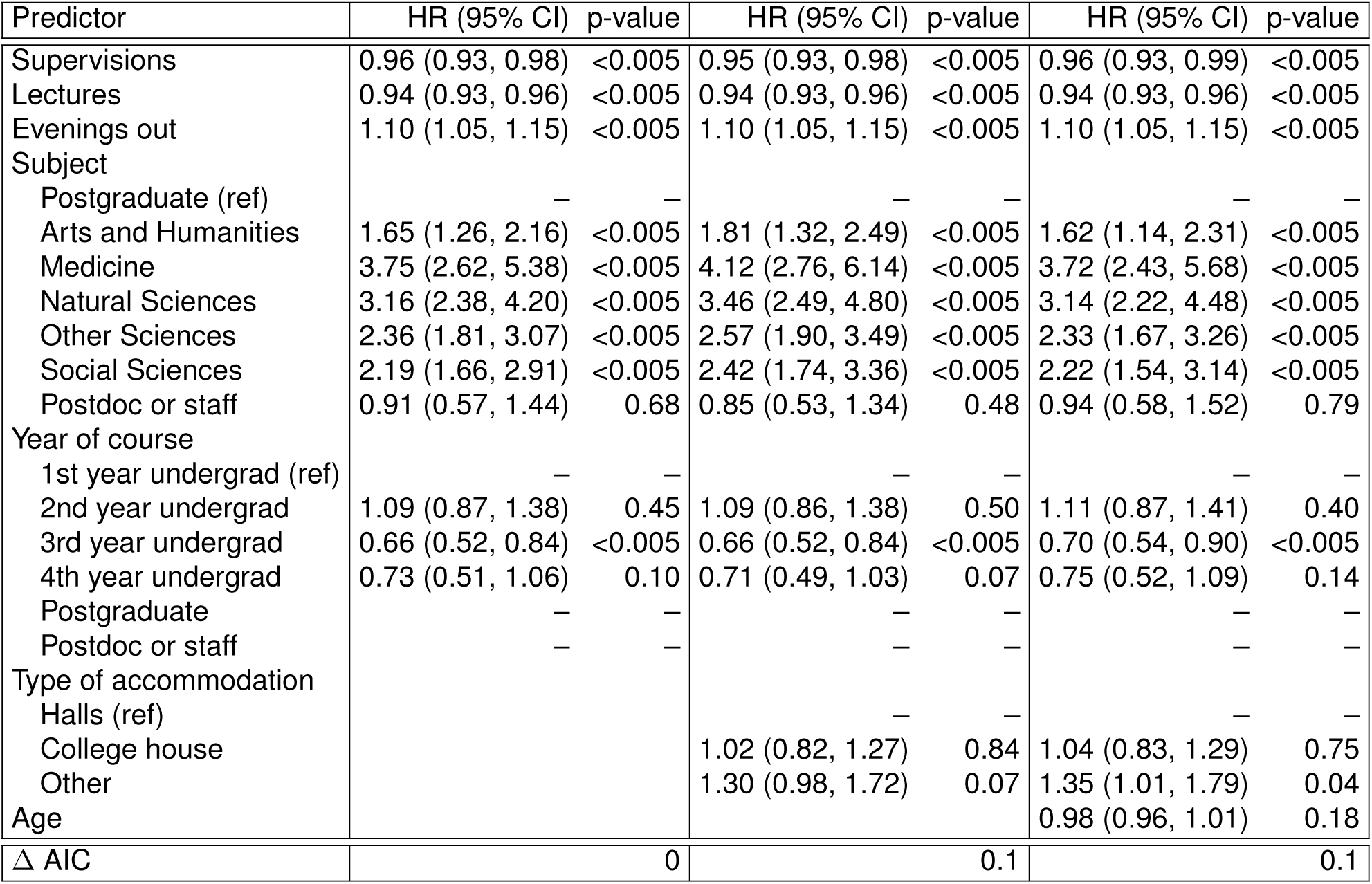
Predictors and hazard ratio (HR) estimates for the three best linear models for symptom onset date during the first term (from 2 October – 30 November 2007) by AIC. The next best model has ΔAIC = 0.8.

All models in Table 2 found that the number of evenings out during the previous week increased the risk of symptom onset by a factor of 10% per evening out. The models also consistently found undergraduates in all subjects to be at higher risk than postgraduates, but this risk varied according to subject (Table 2). Compared to postgraduates, undergraduates studying medicine were found to be at the most risk of symptom onset, about 250-300% more, Natural Sciences undergraduates had a increased risk of over 200%, Social Sciences or Other Sciences undergraduates had a 100-150% increased risk and Arts and Humanities undergraduates had the smallest increase in risk of 60-80%. Although postdoctoral researchers and staff were on average less at risk than postgraduates, there were not enough (21) in the sample to give a significant result.

No significant difference was found between participants living in halls and in college houses, but those not living in college accommodation were found to be around 30% more at risk than those in halls, although this was not highly significant (Table 2). This may also be influenced by interactions with other variables; undergraduates living in halls report infection marginally earlier than those outside college accommodation, but the few postgraduates who live in halls report infection later than those outside college accommodation (Figure S6). Age was also not found to be a significant risk factor in the third proportional hazards model, probably due to correlation with the other predictors.

The variable ‘lectures’ violated the proportional hazards assumption (p-value = 3.5e-15, Table S2). Since this variable recorded the number of lectures in the week previous to symptom onset, students with symptoms in the first week of term reported few lectures from the previous week (Figure 4c), and the lectures coefficient was also seen to be time-dependent in this period (Figure S7). Stratifying the lectures effect between the first few days and the rest of term still violated the proportional hazards assumption but less significantly (p-value = 0.014 for six days, p-value = 0.002 for seven days). The six-day-stratified model found a similar effect for lectures as the time-independent model, but lectures were only significant in the first six-day period (Table S3). Other parameter estimates were broadly consistent, although the increased risks of undergraduate subjects compared to postgraduates were less extreme, and fourth-year undergraduates were not significantly different from first-year undergraduates. Explicitly modelling the time-dependence of the lectures variable instead gives estimates for all other parameters consistent with the time-independent model (Table S3).

### 4.2 Risk ratios for different reporting rates

Raw numbers of reported illness within different student subpopulations were highly dependent on both the subpopulation’s size and could also be dependent on the survey reporting rate. For example, the number of reported cases of symptoms was 36 times more for undergraduates in halls than undergraduates in other accommodation, which was skewed by the majority of the Cambridge undergraduate population living in halls, and could also have been affected by different survey reporting rates between students living in different types of accommodation. Therefore, we explored how reporting rates within different subpopulations can influence risk ratios for this study and give example risk ratios using response rates from other surveys.

Risk ratios depend linearly on the ratio of the subpopulations’ reporting rates, as approximately do the bounds on the region of 95% significance (Figure 8). Hence risk ratios estimated using the unequal response rates from the surveys in Table S1 differed from the risk ratios obtained from assuming equal reporting rates (Table 3). The estimated risk ratio of male and female students depended on the survey used; response rates from [5] and [14] suggested respectively greater risk of symptom onset in male than female students, or an insignificant difference in risk (Figure 8a). However, the response rates from three surveys all estimated first-year undergraduates to have higher risk, between two to three times higher, of symptom onset compared to other undergraduates (Figure 8b). Other estimates from the response rates of the only available survey suggest higher risk of symptom onset in undergraduates compared to postgraduates (over 1.5 times as high; Figure 8c), and in undergraduates living in halls compared to those in other accommodation (over 2 times as high; Figure 8d).

**Table 3:** Risk ratio (RR) estimates and 95% CIs. The second column contains RR estimates assuming equal reporting rates between the subpopulations in the first column, with the specific 95% CI if these equal reporting rates are 1. The third column contains RR estimates and 95% CIs from assuming response rates from other surveys, given in the fourth column. These surveys are further described in Table S1.

| Subpopulations | RR assuming equal reporting rates (95% CI for reporting rates = 1) | RR assuming response rates from other surveys (95% CI) | Other survey used |
| --- | --- | --- | --- |
| Female students, male students (ref) | 1.47 (1.31, 1.65) | 0.68 (0.66, 0.70) | [5] |
|  |  | 1.05 (0.99, 1.11) | [14] |
| First-year undergraduates, other undergraduates (ref) | 2.37 (2.04, 2.75) | 3.37 (3.15, 3.61) | [14] |
|  |  | 2.25 (2.06, 2.46) | [33] (online) |
|  |  | 2.60 (2.37, 2.85) | [33] (paper) |
| Undergraduates, postgraduates (ref) | 1.34 (1.18, 1.53) | 1.56 (1.45, 1.68) | [14] |
| Undergraduates living in halls, undergraduates living elsewhere (ref) | 3.34 (2.15, 5.19) | 2.15 (1.73, 2.67) | [14] |

**Figure 8:**
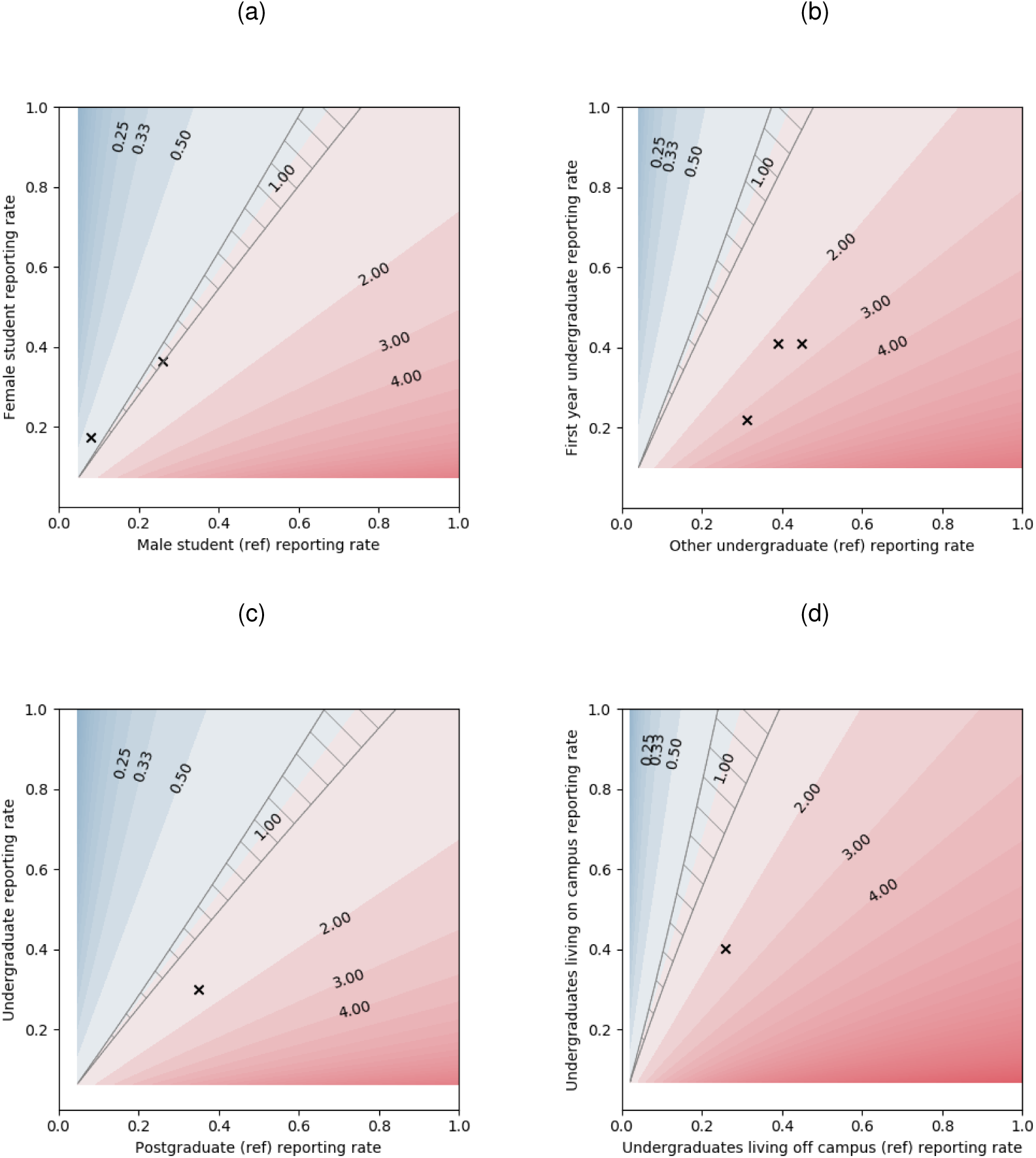
Risk ratio (RR) contours for different reporting rates, with contours RR = 1/4, 1/3, 1/2, 1, 2, 3, 4 labelled. Red areas have RR>1, where the y-axis subpopulation has more risk of symptom onset than the x-axis reference subpopulation. Blue areas have RR<1, where the y-axis subpopulation has less risk than the x-axis reference subpopulation. Inside the hatched area, the RR is not significant, i.e. the 95% CI includes 1 and there is no significant risk difference between the two subpopulations. The RR estimates obtained using response rates from other surveys are plotted as crosses, and the values are given in Table 3.

## 5 Discussion

In this paper, we have shared and described the data from a UK university survey on self-reported coughs, colds and flu-like symptoms and lifestyle factors from 2007-2008. The ongoing COVID-19 pandemic has motivated our retrospective analysis and dissemination of findings to help understand transmission of respiratory infection amongst university populations.

Overall, the data from this study shows some basis for the term ‘*freshers’ flu*’ as risk ratio estimates suggest first-year undergraduates had an overall higher risk than other undergraduate years (Table 3), also seen in other studies [9, 10], but our multivariate analysis found first-year undergraduates were only significantly more at risk over time and hence more likely to report infection earlier than third-year undergraduates, and not second or fourth years. These findings could have been confounded by reporting rate if perhaps freshers were more likely to sign up for a survey on ‘*freshers’ flu*’. The raw data and analysis also suggested undergraduates were more at risk of symptom onset and reporting illness earlier than postgraduates (Figures 3b, 3f, Table 2, Table 3), agreeing with the higher undergraduate incidences seen in [7, 6]. Further, there was some evidence that this effect was more extreme for undergraduates studying medicine or any science, but this could instead reflect different reporting rates for subjects.

The multivariate regression implies that the social environment on evenings out may be important for disease transmission and increase risk of ILI, whereas lectures and supervisions do not. It also provides a weak indication that more lectures and supervisions could decrease students’ risk of illness, speculatively this could be by preventing students from engaging in other behaviour that does advance transmission. Incidence differences between students living on campus and off campus vary between studies [7, 14], and in our study, we saw mixed effects for undergraduates, postgraduates and overall (Figure S6, Table 2).

Attack rates cannot be determined solely from this survey as only people who reported symptoms were recruited, and neither can a comparison be made between the behaviour of those who were and were not infected. Comparing the demographics of the dataset to the demographics of the university population and using subpopulation survey response rates from other surveys of university students can give an indication of actual incidence between subpopulations (Figure 8), since survey response rate has been observed to vary by individual factors such as gender [33]. However, survey response rates can also vary by institution and the method of survey recruitment [33], as well as perhaps by country. Assuming equal response rates between those with and without symptoms might also not be valid, as having symptoms could conceivably make someone more motivated to contribute to the survey, or less if they feel ill and perhaps did not see the survey advertised. Furthermore, risk ratios and proportional hazards models are not built for the contagious nature of infectious diseases and the possible resulting dependence between individuals [34].

Other limitations also come with self-selected surveys. Sampling biases make it hard to separate epidemiological clusters from clusters of related people responding to the survey. Although consistent advertising of the survey was attempted, biases could still occur from location of posters and advertisement by undergraduate and postgraduate common rooms or college nurses. In addition, most participants reported a recent symptom onset to the survey (Figure S2), so patterns of reported infection could simply reflect the pattern of interest and awareness of the study, which is likely to decline over time. Self-reporting in this study could also have led to inaccurate data, as participants reported information about symptom onset up to four months previously (Figure S2) and there were no interactions with those who participated beyond completion of the online survey. We also note that the ability of self-defined coughs, colds and flu-like symptoms to predict actual infectious illness levels is unknown, as the symptoms are not well-defined and can be caused by many types of infections, or indeed non-infectious causes. Despite the vague biological criteria, clear term-time epidemic-like waves are seen in the data, with the first wave peaking before the middle of the first term (although this could again be merely illustrating the wave of interest in the study across terms).

Despite all the caveats, this exploratory study is one of few recent datasets about infectious disease and in a university prior to 2020 [11, 14, 15, 12], and is still, as far as we are aware, the most detailed survey of its kind in a UK university. This pilot study shows that an inexpensive and relatively simple survey to run can receive a large response (over 5% of the university’s student population) and pick up interesting signals. For future surveys, recruiting via email, including those without symptoms, or regular surveys like with *Flusurvey* [35] would allow response rates and incidences to be calculated more easily, but some previously mentioned biases would still remain. However, while more complex studies could be planned, there is huge value in running similar studies to this one that are relatively easy and quick to arrange and execute. Doing this in more universities in the future can help to gather the data needed to answer the pertinent questions of infectious disease transmission and control within universities.

## Data Availability

The anonymous and non-identifiable dataset from this study is available to be requested from the corresponding author.

## Author contributions

**Conceptualisation:** KTDE. **Methodology:** KTDE, MLT. **Software:** MLT. **Formal analysis:** KTDE, MLT. **Investigation:** KTDE, MLT. **Data Curation:** KTDE, MLT. **Supervision:** JRG. **Validation:** KTDE, MLT, EMH, MJT, JMR, MJK, JRG. **Visualisation:** MLT. **Writing - Original Draft:** MLT. **Writing - Review & Editing:** KTDE, MLT, EMH, MJT, JMR, MJK, JRG.

## Data availability

The anonymous and non-identifiable dataset from this study may be requested from the corresponding author.

