## Supplemental information for "Coughs, Colds and “Freshers’ Flu” Survey in the University of Cambridge, 2007-2008"

### Supplementary information

#### Supplementary data S1

Data from the survey, with variable names as described in Table 1. This supporting data is not allowed on the preprint server; readers may contact the corresponding author to request this data.

#### Supplementary text S1: dataset changes for non-identifiability purposes

Some variables in the dataset were amended to preserve non-identifiability:

- **Subject:** This variable originally specified degree course and was aggregated into broader groups of departments. For undergraduates, aggregation was based on the undergraduate courses covered by the university's School of Humanities and Social Sciences (Economics, Education Studies, History, Social & Political Sciences, Archaeology & Anthropology, Law, Land Economy), School of Arts and Humanities (Architecture, History of Art, Oriental Studies, Classics, Theology & Religious Studies, English, Modern & Medieval Languages, Music, Philosophy, Anglo-Saxon, Norse & Celtic, Linguistics), School of Clinical Medicine (Medicine and Veterinary Medicine), the Natural Sciences degree course, and the remaining science courses in the School of Technology and the School of Physical Sciences (Engineering, Chemical Engineering, Computer Science, Management Studies, Mathematics, Geography). For non-undergraduates, the 'academic status' variable was duplicated, as the subject question was not answered consistently, by postgraduates for example.

- **Number of Cambridge Facebook friends:** Only numeric answers were retained for this originally free-text variable.

- **University department:** This variable originally had free text answers, so was removed from the dataset.

- **Other behavioural changes experienced:** This variable originally had free text answers, so was removed from the dataset.

#### Supplementary tables and figures

| | Survey description | Population | Year |
|---|---|---|---|
| [1] | Online survey on ILIs and impact with monthly follow-ups (we use data for initial survey only), recruited by email | Students at the University of Minnesota, Twin Cities campus | 2002-2003 |
| [2] | Online survey on ILIs and related health information, recruited by email | University of Delaware students | 2009 |
| [3] | The National Survey of Student Engagement, online survey recruited by email or paper survey by mail | Students from 91 US universities (for online survey) and 226 US universities (paper survey) - response rates averaged across students | 2002-2003 |

Table S1: The surveys from which survey response rates are used to estimate odds ratios in Table 3

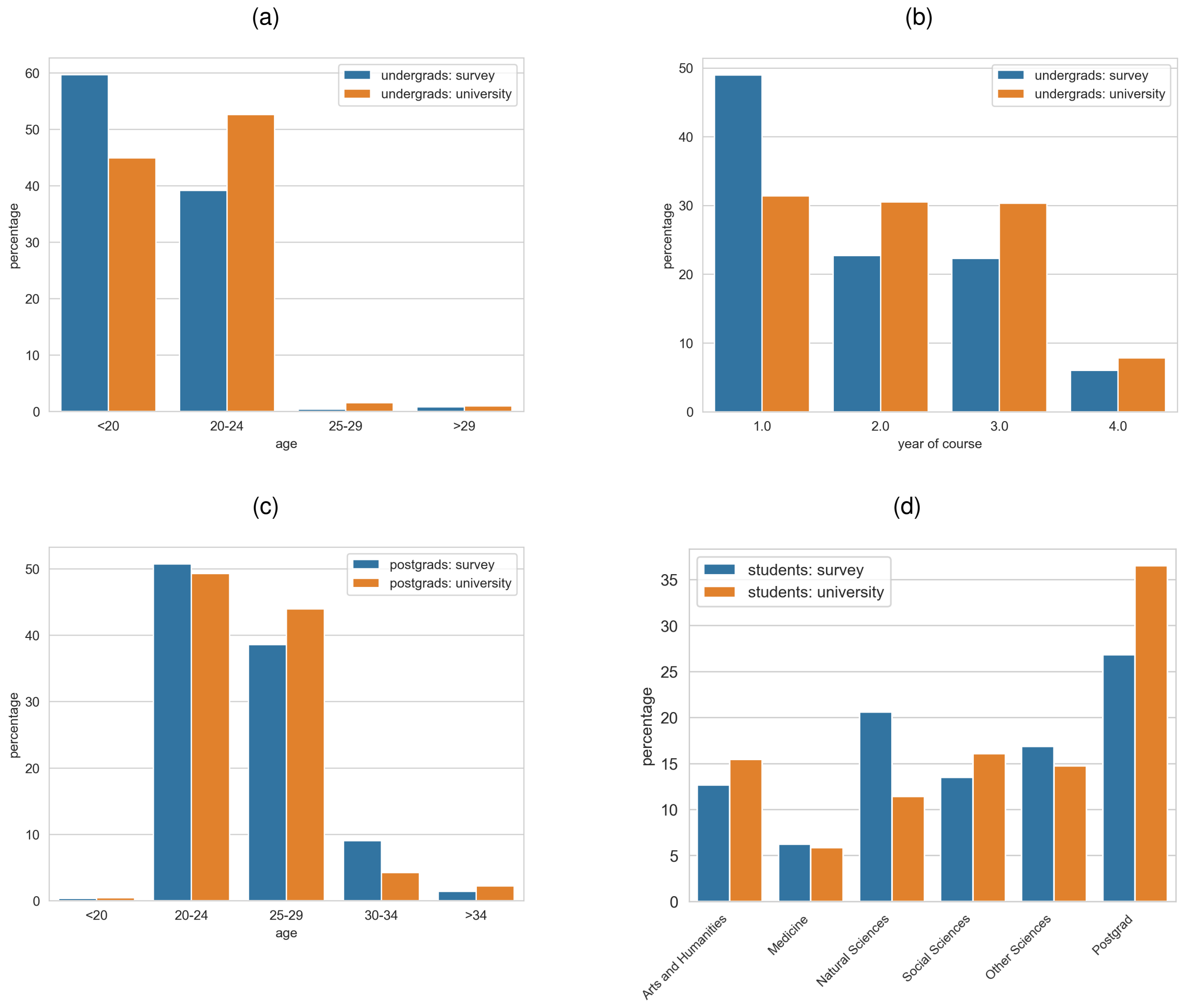


Figure S1: Distribution of undergraduates in the survey compared to the undergraduate population by (a) age and (b) year of course. (c) Distribution of postgraduates in the survey by age, compared to the postgraduate population. (e) Distribution of undergraduate subject and postgraduate numbers in the survey compared to the student population. Student population statistics are of full-time undergraduate or postgraduate students in the academic year 2007-2008 from [4].

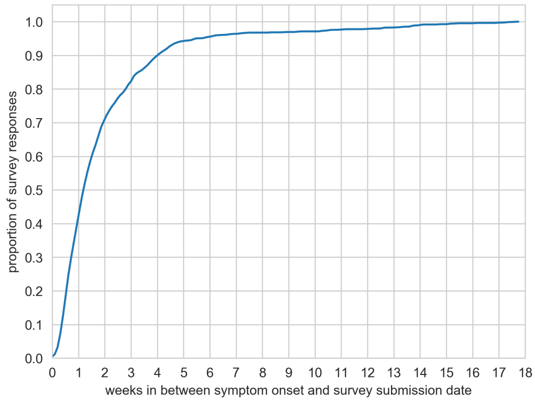


Figure S2: Cumulative distribution of number of weeks between survey submission date and reported symptom onset date.

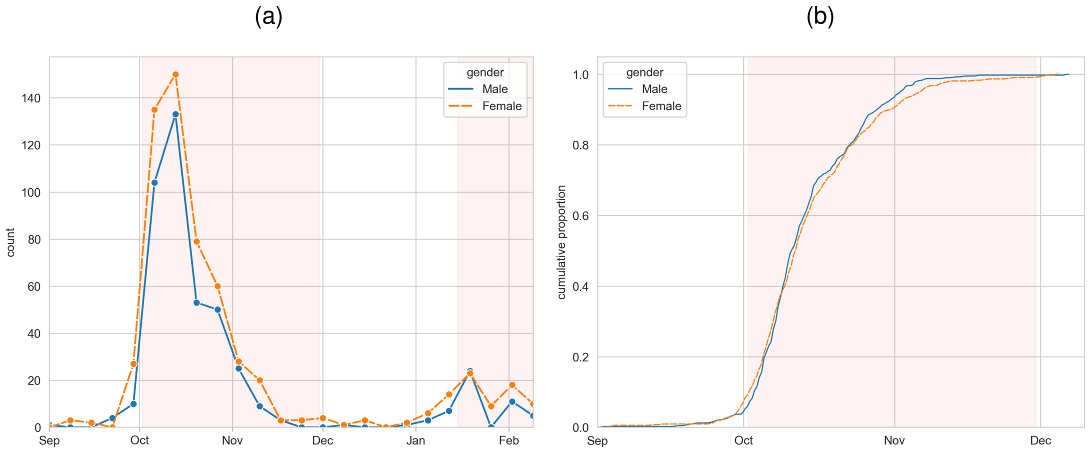


Figure S3: (a) Distribution of weekly reported cases and (b) cumulative distribution of reported cases over the first wave by gender. University of Cambridge academic terms are highlighted in orange.

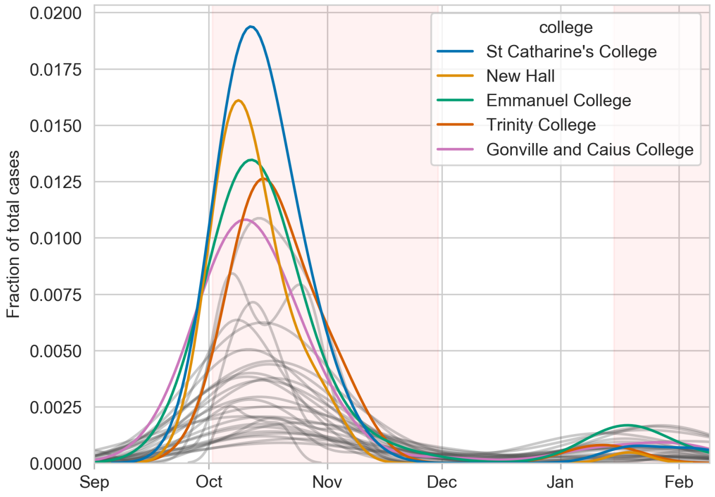


Figure S4: Smoothed distributions of reported cases by symptom onset date and college. The five colleges with the most reported cases are in colour. Distributions are smoothed using Gaussian kernels. University of Cambridge academic terms are highlighted in orange.

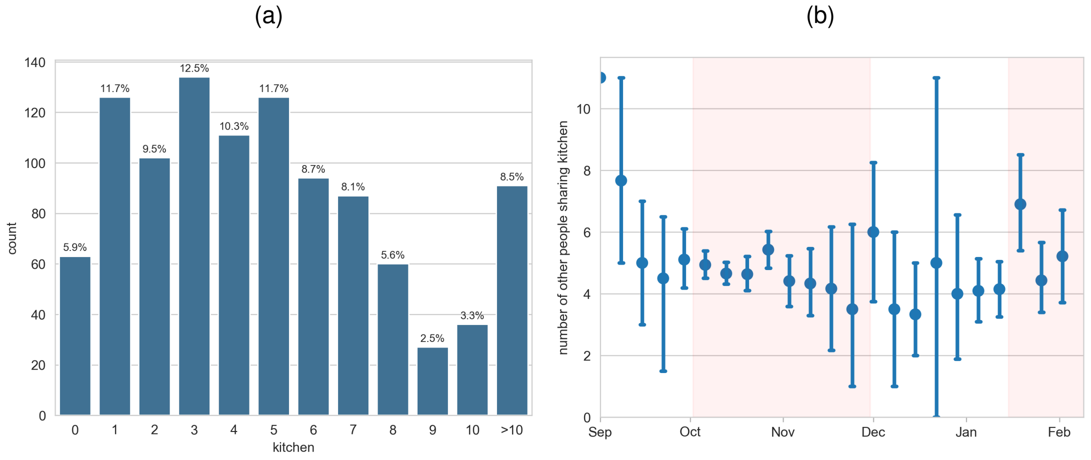


Figure S5: (a) Distribution of number of other people sharing kitchen facilities. (b) Mean number of other people sharing kitchen facilities distributed by date of symptom onset, with standard error. University of Cambridge academic terms are highlighted in orange.

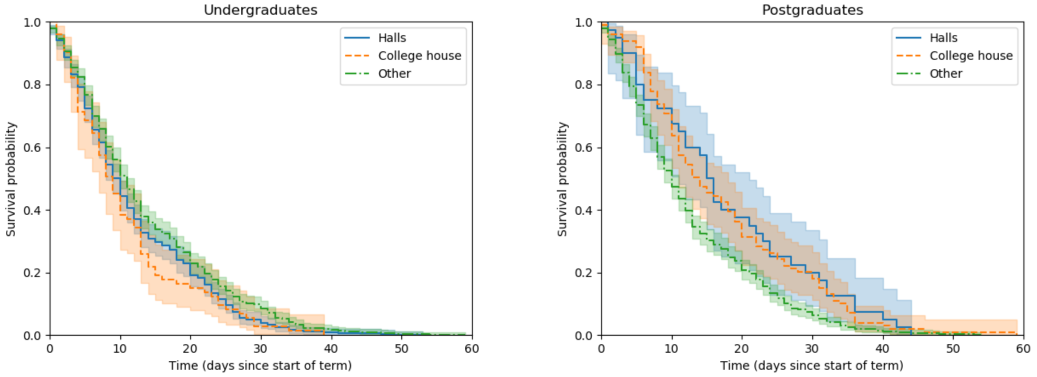


Figure S6: Kaplan-Meier survival curves with 95% CIs for undergraduates and postgraduates by type of accommodation

| Predictor | p-value |
|---|---|
| Supervisions | 0.20 |
| Lectures | 3.5e-15 |
| Evenings out | 0.32 |
| Subject | 0.67 |
| Year of course | 0.15 |

Table S2: P-values from the Grambsch-Therneau proportional hazards assumption test [5] for the best model in Table 2.

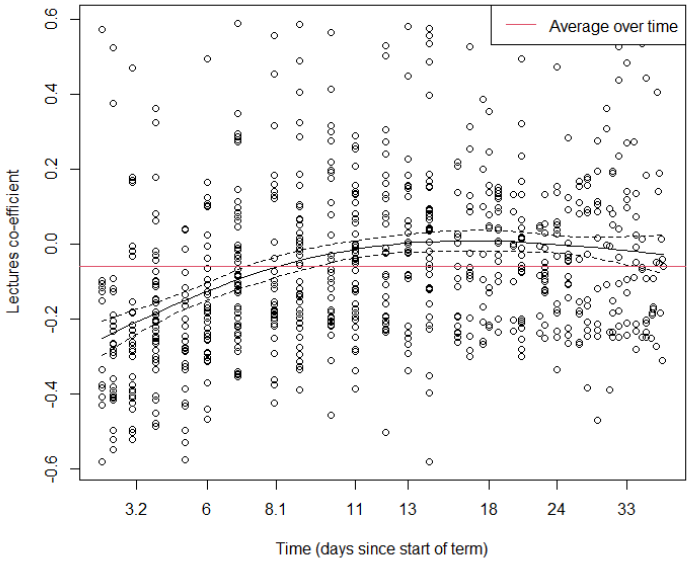


Figure S7: Time-dependent co-efficient estimate with 95% CI for lectures variable in a proportional hazards model with the same predictors as the best model in Table 2.

| | Time-independent model | | Time-stratified model | | Time-dependent model | |
|---|---|---|---|---|---|---|
| Predictor | HR (95% CI) | p-value | HR (95% CI) | p-value | HR (95% CI) | p-value |
| Supervisions | 0.96 (0.93, 0.98) | <0.005 | 0.96 (0.94, 0.99) | 0.01 | 0.96 (0.93, 0.98) | <0.005 |
| Lectures | 0.94 (0.93, 0.96) | <0.005 | – | – | time-dependent | |
| Lectures ($t \leq 6$) | | | 0.93 (0.89, 0.96) | <0.005 | | |
| Lectures ($t \geq 7$) | | | 0.99 (0.97, 1.00) | 0.07 | | |
| Evenings out | 1.10 (1.05, 1.15) | <0.005 | 1.07 (1.02, 1.12) | <0.005 | 1.10 (1.05, 1.15) | <0.005 |
| Subject | | | | | | |
| Postgraduate (ref) | – | – | – | – | – | – |
| Arts and Humanities | 1.65 (1.26, 2.16) | <0.005 | 1.39 (1.06, 1.83) | 0.02 | 1.60 (1.22, 2.11) | <0.005 |
| Medicine | 3.75 (2.62, 5.38) | <0.005 | 1.78 (1.25, 2.53) | <0.005 | 3.25 (2.31, 4.57) | <0.005 |
| Natural Sciences | 3.16 (2.38, 4.20) | <0.005 | 1.80 (1.37, 2.37) | <0.005 | 3.03 (2.35, 3.90) | <0.005 |
| Other Sciences | 2.36 (1.81, 3.07) | <0.005 | 1.55 (1.19, 2.02) | <0.005 | 2.23 (1.73, 2.86) | <0.005 |
| Social Sciences | 2.19 (1.66, 2.91) | <0.005 | 1.66 (1.26, 2.20) | <0.005 | 2.09 (1.58, 2.75) | <0.005 |
| Postdoc or staff | 0.91 (0.57, 1.44) | 0.68 | 0.94 (0.59, 1.49) | 0.79 | 0.92 (0.58, 1.46) | 0.59 |
| Year of course | | | | | | |
| 1st year undergrad (ref) | – | – | – | – | – | – |
| 2nd year undergrad | 1.09 (0.87, 1.38) | 0.45 | 1.10 (0.87, 1.38) | 0.44 | 1.06 (0.84, 1.34) | 0.45 |
| 3rd year undergrad | 0.66 (0.52, 0.84) | <0.005 | 0.71 (0.55, 0.9) | <0.005 | 0.66 (0.52, 0.83) | <0.005 |
| 4th year undergrad | 0.73 (0.51, 1.06) | 0.1 | 1.02 (0.71, 1.48) | 0.9 | 0.75 (0.52, 1.08) | 0.01 |
| Postgraduate | – | – | – | – | – | – |
| Postdoc or staff | – | – | – | – | – | – |

Table S3: Predictors and hazard ratio (HR) estimates for proportional hazards models for symptom onset date during the first term (from 2 October – 30 November 2007). The three models shown fit the lectures predictor respectively as time-independent (same as best model in Table 2), time-stratified, and time-dependent.
